# Three Risk Scores For Mortality Prediction In Minimally Invasive Cardiac Surgery

**DOI:** 10.1101/2020.06.03.20120949

**Authors:** R. Margaryan, G. Bianchi, G. Concistre, T. Gasbarri, E. Kallushi, P. Farneti, M. Solinas

## Abstract

**Objective:** The Society of Thoracic Surgeons score performance relative to other scores in minimally invasive cardiac surgery is not known.

**Methods:** Patients who underwent surgery from 2003 to 2018 identified from database. Additional variables included for STS score calculation, EuroSCORE II and age, creatinine and ejection fraction score calculation.

**Results:** A total of 4751 patients were identified from main database. There were actual 47 (0.99%) hospitals deaths. The mean STS score predicted mortality were 2.0 ± 2.1. Discriminatory power was uniformly good (for STS Mortality: area under curve, 0.86; 95% confidence interval, 0.81 - 0.91). The mean EuroSCORE II predicted mortality were 2.9 ± 3.8. Discriminatory power was uniformly good similar to that of STS (for EuroSCORE II Mortality: area under curve, 0.9; 95% confidence interval, 0.86 - 0.93). The mean ACEF predicted mortality were 2.5 ± 2.3. Discriminatory power was uniformly good but inferior to that of STS and EuroSCORE II (for ACEF Mortality: area under curve, 0.72; 95% confidence interval, 0.65 - 0.8).

Calibration pattern for STS score was the best for of mortality prediction (p < 0.01), EuroSCORE II and ACEF were constantly overestimating mortality (respectively, p < 0.01 and p < 0.01). scores.

**Conclusions:** The STS score has acceptable discrimination power for this sub-population. However, it is not calibrated for the the subset. EuroSCORE II is has good discrimination power, but not calibrate for the this subset of patients. ACEF score had similar performance to EuroSCORE. No algorithm seems suitable for accurate risk estimation.

## Introduction

The European System for Cardiac Operative Risk Evaluation (EuroSCORE) has two editions and the last update was committed in 2012^1^. It was a substitution for older EuroSCORE developed originally in 1991^2^. These older versions and new version gained wide popularity and are used worldwide for cardiac surgical services^3-5^. They were tested and validated for by different groups worldwide^4^. Moreover, additive/logistic EuroSCORE and new EuroSCORE II have been employed in recent years together with other evaluation scores, for example Society of Thoracic Surgeons (STS) score and age, creatinine and ejection fraction score (ACEF), for the screening, selection of high risk patients for hybrid surgical techniques, for instance, TAVI^6^, but analysis of its performance within these specific surgical sub-populations has first underlined tendency to over-predict the risk of mortality and morbidity. Barili et al^7^ showed that ACEF score in predicting in-hospital mortality in elective and non-elective cardiac surgery comparable to EuroSCORE II. Nonetheless, it is not as satisfactory as the new EuroSCORE II, as its discrimination is significantly lower and it is also mis-calibrated^8^. STS score performs with high predictive ability in different population^5^ but it seems that it is not well calibrated for other then american populations. However, there are no reports which have performed external calibration of STS score, ACEF score comparable to EuroSCORE II.

### Objectives

We sought to analyse STS, ACEF and EuroSCORE II performance in MICS European population (Massa experience).

## Material and Methods

### Study Design and Participants

The study population included all patients who underwent minimally invasive cardiac surgery (MICS) in 15 years period (from 2003 to 2018 Sep 1, overall 4751 patients enrolled) within the department of single center. They all underwent MICS procedure (see Table 1). Trans-catheter/per-cutaneous valve implantation procedures were excluded from the study group. However, we kept trans apical aortic valve implantations via left minithoracotomy (TATAVI) Preoperative and demographic information, operative data and peri-operative mortality, and complication for tall patients were retrieved from institutional databases that were prospectively collected (see Table 1. The Institutional Ethical Committee approved the study and the requirement for informed written consent was waived on the condition that subjects’ identities were masked. For the evaluation and validation of the performance of the three scores were calculated for each patient according to published guidelines with a dedicated software.

**Table 1:** Summary descriptives table by groups of ‘Sex’

|  | F<br>N=2152 | M<br>N=2599 | p.overall |
| --- | --- | --- | --- |
| Age | 69.3 (12.2) | 65.6 (12.7) | <0.001 |
| BSA | 1.71 (0.80) | 1.92 (0.25) | <0.001 |
| BMI | 25.8 (4.73) | 26.2 (3.72) | 0.003 |
| LV_EF | 57.4 (8.07) | 56.2 (8.82) | <0.001 |
| hispanic_ethnicity: No | 2152 (100%) | 2599 (100%) | . |
| NYHA: |  |  | <0.001 |
| I | 129 (7.38%) | 377 (18.8%) |  |
| II | 913 (52.3%) | 1095 (54.6%) |  |
| III | 637 (36.5%) | 474 (23.7%) |  |
| IV | 68 (3.89%) | 58 (2.89%) |  |
| Procedure_Type: |  |  | 0.012 |
| Elective | 1798 (83.6%) | 2093 (80.5%) |  |
| Urgent | 10 (0.46%) | 13 (0.50%) |  |
| Emergent | 2 (0.09%) | 0 (0.00%) |  |
| Salvage | 342 (15.9%) | 493 (19.0%) |  |
| Procedure_Grouping: |  |  | <0.001 |
| AVP | 761 (35.4%) | 832 (32.0%) |  |
| MIDCAB | 58 (2.70%) | 284 (10.9%) |  |
| MVP | 1184 (55.0%) | 1274 (49.0%) |  |
| OTHER | 114 (5.30%) | 157 (6.04%) |  |
| TATAVI | 35 (1.63%) | 52 (2.00%) |  |
| Renal_Insufficiency: |  |  | 0.015 |
| No | 2114 (98.2%) | 2519 (96.9%) |  |
| Yes(No Dialysis) | 29 (1.35%) | 59 (2.27%) |  |
| Yes(Dialysis) | 9 (0.42%) | 21 (0.81%) |  |

### Statistical methods

The performance of the all prediction models was analysed focusing on discrimination power and calibration^9,10^. The discrimination performance indicates the extent to which the model distinguishes between patients who will die or survive in the peri-operative period. It was evaluated by constructing receiver operating characteristic curves for each model and calculating the area under the curve (AUC) with with 95% confidence intervals^11,12^. The comparison among the curves was analysed with Delong, bootstrap and Venkatraman methods^12^. Another index used to evaluate the predictive abilities was Somers’ *D_xy_* rank correlation between predicted probabilities and observed responses. When *D_xy_* = 0, the model is making random prediction, when *D_xy_* = 1, the predictions are perfectly discriminating.

Calibration refers to the agreement between observed and predicted outcomes. The calibration performance can be evaluated by generating calibration plots that visually compare the prediction with the observed probability.^9,12,13^ Agreement k statistics was expressed in plain English as follows: 0 - 0.20 slight; 0.21 - 0.40 fair; 0.41 - 0.60 moderate; 0.61 - 0.80 substantial; 0.81 - 1.00 almost perfect.^14^ The calibration was tested with the Hosmer-Lemeshow goodness-of-fit test, which compares observed to predicted values by decile of predicted probability. The accuracy of the model was also tested calculating the Brier score.^12,13^ Missing values were substituted by means of multiple imputation in order to reduce bias and increase statistical power^15^. Two sided statistics were performed with a significance level of 0.05. For all analysis R Statistical Computing Environment^16^ were used with RStudio (RStudio (2018). RStudio: Integrated development environment for R (Version 1.1.442) [Computer software]. Boston, MA. Retrieved May 1, 2018. Available from http://www.rstudio.org/)

## Results

### Participants and Descriptive Data

There were 47 (0.99%) hospitals deaths. The mean ± SD values of STS, EuroSCORE II and ACEF of population were 2.0 ± 2.1, 2.9 ± 3.8 and 2.5 ± 2.3, respectively (see Table 2). Baseline characteristics are described in Table 1.

**Table 2:** Summary descriptives table by groups of ‘Type’

|  | AVP<br>N=1593 | MIDCAB<br>N=342 | MVP<br>N=2458 | OTHER<br>N=271 | TATAVI<br>N=87 | p.overall |
| --- | --- | --- | --- | --- | --- | --- |
| STS_Mortality | 2.19 (1.77) | 1.44 (2.31) | 1.88 (2.18) | 1.86 (1.54) | 5.35 (3.30) | <0.001 |
| EuroSCORE_II | 2.53 (2.72) | 3.41 (3.71) | 2.82 (4.06) | 3.49 (3.74) | 8.09 (7.02) | <0.001 |
| ACEF | 2.89 (2.28) | 2.90 (2.59) | 2.24 (2.16) | 1.85 (1.42) | 5.56 (4.06) | <0.001 |
| Observed_Mortality: |  |  |  |  |  | 0.004 |
| No | 1583 (99.4%) | 339 (99.1%) | 2433 (99.0%) | 267 (98.5%) | 82 (94.3%) |  |
| Yes | 10 (0.63%) | 3 (0.88%) | 25 (1.02%) | 4 (1.48%) | 5 (5.75%) |  |

**Table 3:** Model calibration statistic

|  | STS | EuroSCORE II | ACEF |
| --- | --- | --- | --- |
| Somers' D~xy~ rank correlation | 0.73 | 0.79 | 0.44 |
| ROC area (C Statistic) | 0.86 | 0.90 | 0.72 |
| Unreliability index U | 0.01 | 0.02 | 0.01 |
| Unreliability index U P-value | 0.00 | 0.00 | 0.00 |
| Brier score | 0.01 | 0.01 | 0.01 |
| Spiegelhalter Z-test for calibration accuracy | 0.02 | 0.03 | 0.03 |
| Spiegelhalter Z-test two-tailed P-value | 0.01 | 0.02 | 0.02 |

### Performance of the scores

STS discriminatory power was uniformly good (for STS Mortality: area under curve, 0.86; 95% confidence interval, 0.81 - 0.91, see Figure 1 A). EuroSCORE II discriminatory power was uniformly good similar to that of STS (for EuroSCORE II Mortality: area under curve, 0.9; 95% confidence interval, 0.86 - 0.93). ACEF discriminatory power was uniformly sufficient but inferior to that of STS and EuroSCORE II (for ACEF Mortality: area under curve, 0.72; 95% confidence interval, 0.65 - 0.8). Calibration pattern for STS score was underestimating mortality after the limit of available data and showed scarce fit (χ^2^ = 34.29,p < 0.01). However in the range of 0-20 % it showed almost perfect calibration (see Figure 1 A). EuroSCORE II and ACEF were overestimating mortality constantly and also showed bad fit (respectively, χ^2^ = 69.27, p < 0.01 and χ^2^ = 49.57, p < 0.01), and showed not good calibration (see Figure 1 B, C).

**Figure 1:**
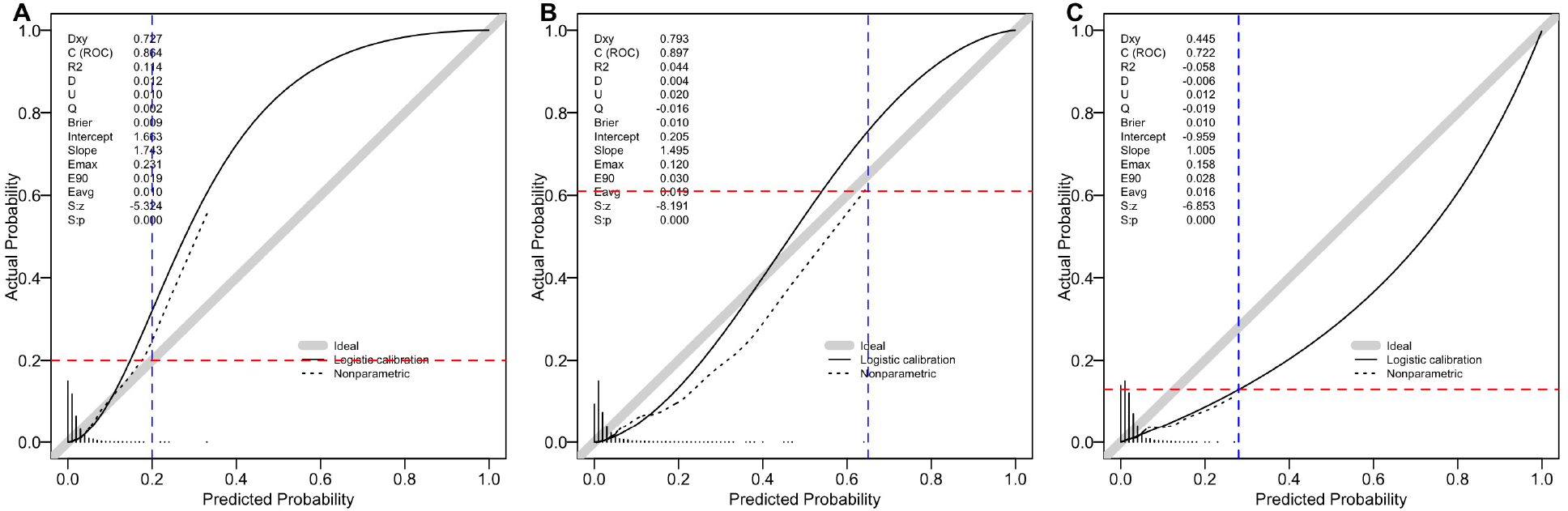
The calibration curves of STS (A), EuroSCORE II (B) and ACEF (C) scores.

**Figure 2:**
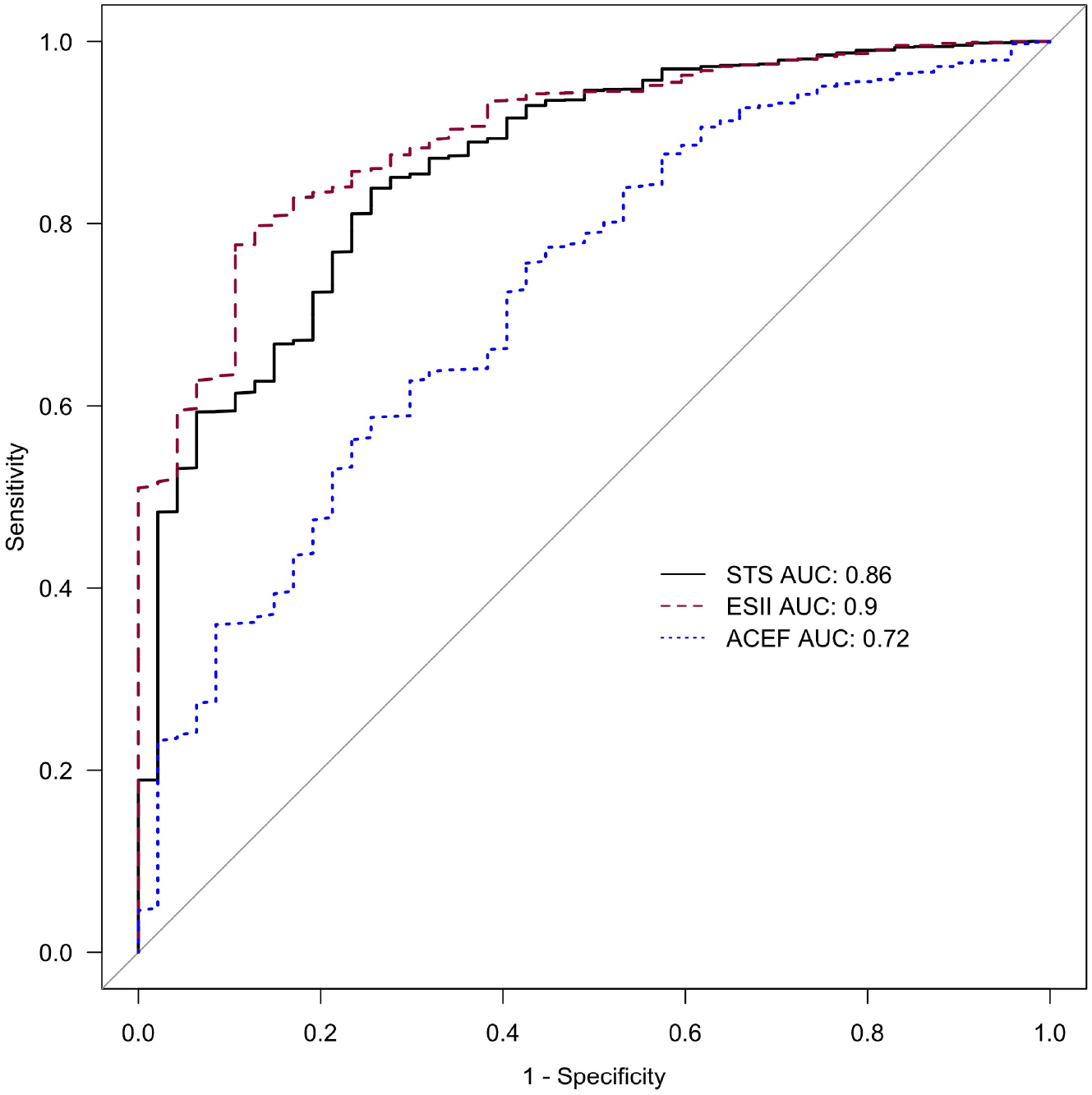
Area Under The Curve (AUC) of Receiver Operating Characteristics (ROC) curves for thre scores)

**Figure 3:**
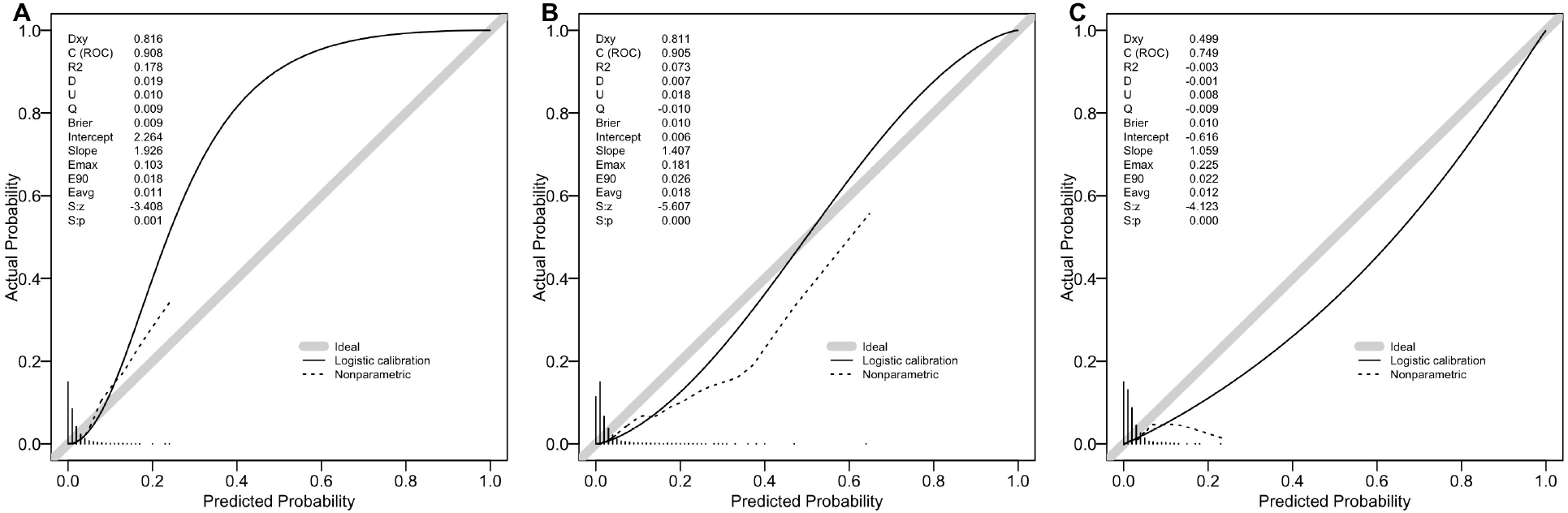
Calibration for the subgroup of MVP(isolated mitral valve procedures)

## Subgroup Performances

### Aortic Valve Procedures

STS discriminatory power was uniformly good (for STS Mortality: area under curve, 0.83; 95% confidence interval, 0.7 - 0.96). Calibration pattern for STS score was similar to all cohort calibration (χ^2^ = 21.6,p < 0.01). EuroSCORE II discriminatory power was uniformly good similar to that of STS (for EuroSCORE II Mortality: area under curve, 0.87; 95% confidence interval, 0.75 - 1). ACEF discriminatory power was uniformly sufficient but inferior to that of STS and EuroSCORE II (for ACEF Mortality: area under curve, 0.72; 95% confidence interval, 0.55 - 0.89, see Table 2). EuroSCORE II and ACEF were overestimating mortality similar to main cohort (respectively, χ^2^ = 25.68, p < 0.01 and χ^2^ = 31.47, p < 0.01).

### Mitral Valve Procedures

STS discriminatory power was uniformly good (for STS Mortality: area under curve, 0.91; 95% confidence interval, 0.85 - 0.97). Calibration pattern for STS score was similar to all cohort calibration (χ^2^ = 18.48,p = 0.02). EuroSCORE II discriminatory power was uniformly good similar to that of STS (for EuroSCORE II Mortality: area under curve, 0.91; 95% confidence interval, 0.86 - 0.95). ACEF discriminatory power was uniformly sufficient but inferior to that of STS and EuroSCORE II (for ACEF Mortality: area under curve, 0.75; 95% confidence interval, 0.66 - 0.84, see Table 2). EuroSCORE II and ACEF were overestimating mortality similar to main cohort (respectively, χ^2^ = 34.42, p < 0.01 and χ^2^ = 18.43, p = 0.02).

### Mitral Valve Procedures

STS discriminatory power was uniformly good (for STS Mortality: area under curve, 0.91; 95% confidence interval, 0.85 - 0.97). Calibration pattern for STS score was similar to all cohort calibration (χ^2^ = 18.48,p = 0.02). EuroSCORE II discriminatory power was uniformly good similar to that of STS (for EuroSCORE II Mortality: area under curve, 0.91; 95% confidence interval, 0.86 - 0.95). ACEF discriminatory power was uniformly sufficient but inferior to that of STS and EuroSCORE II (for ACEF Mortality: area under curve, 0.75; 95% confidence interval, 0.66 - 0.84, see Table 2). EuroSCORE II and ACEF were overestimating mortality similar to main cohort (respectively, χ^2^ = 34.42, p < 0.01 and χ^2^ = 18.43, p = 0.02).

### Mitral Valve Procedures

STS discriminatory power was uniformly good (for STS Mortality: area under curve, 0.91; 95% confidence interval, 0.85 - 0.97). Calibration pattern for STS score was similar to all cohort calibration (χ^2^ = 18.48,p = 0.02). EuroSCORE II discriminatory power was uniformly good similar to that of STS (for EuroSCORE II Mortality: area under curve, 0.91; 95% confidence interval, 0.86 - 0.95). ACEF discriminatory power was uniformly sufficient but inferior to that of STS and EuroSCORE II (for ACEF Mortality: area under curve, 0.75; 95% confidence interval, 0.66 - 0.84, see Table 2). EuroSCORE II and ACEF were overestimating mortality similar to main cohort (respectively, χ^2^ = 34.42, p < 0.01 and χ^2^ = 18.43, p = 0.02).

## Discussion

### Key Results

We have demonstrated that STS score in the first part seems to have good discrimination power, however in high risks underestimates the observed mortality, whereas EuroSCORE II and ACEF over-predict the operator risks for MICS over all range of available data. STS seems most calibrated score in a rage of 0 - 20 % of risk. Risk stratification and risk scoring systems in adult cardiac surgery are becoming increasingly important and should have its clinical application when possible. They provide reliable estimation of the risk associated with surgical procedures, permit comparison of outcomes among institutions and surgeons, and may provide a more accurate assessment of the indication for surgery in individual patients by potential risks and benefits.^8,17,18^

In recent years, STS and EuroSCORE II model have been widely used as risk prediction tools in adult cardiac surgery, particularly in European countries. STS recently added to new miocardial revascularisation guideline which underlines its quality and predictive capability^19^. By far, mentioned scores were developed using big data collections^18^. Our group had demonstrated that EuroSCORE II was the best alternative available when comparing to old version and to additive model^8^. There are no reports on how STS score works in selected MICS population. This report is unique in providing an insight how might STS work for minimally invasive cardiac surgery.

Limitations of original EuroSCORE models’ performance were also highlighted by the comparison with the STS score, which predicts more accurately the observed morality, especially in the highest-risk patients. The main reason for that is that the STS score is derived from a much larger data set (with respect to original EuroSCORE and EuroSCORE II) of patients operated in a more current era, and risk models were separately developed in the different surgical categories (CABG, valves, CABG and Valves) and contains more covariates^18^. The EuroSCORE II was mainly conceived to overcome the constant high-grade over-prediction of original EuroSCORE that was present in the literature^20^. In this study particularly we have demonstrated that EuroSCORE has

Our study confirmed the unsatisfactory calibration of EuroSCORE II and ACEF (Figures 1B, C). The STS has after all the best almost perfect calibration until 20 % predicted probability (see Figure 1A), whereas it progressively under-predicts afterward, leading to a calibration to remain distant from ideal calibration line (Figure 1A, diagonal line).

Moreover, EuroSCORE II and ACEF show a somehow constant over-prediction in whole population. This pattern is reported by other authors^4,8^. To our knowledge, there are no other studies that performed external calibration of all three scores. The EuroSCORE II population contained approximately 47% isolated CABG, and hence, score could be more precise in validation of that kind of external population. Barili et al^4^ performed EuroSCORE calibration on isolated CABG data set and found that it had very good calibration but still suffering from poor reliability.

### Limitations

The potential limitation of the current study is its retrospective nature, although data were prospectively collected. All the score contain very small percent of MICS. Because the observed mortality is low, observations number is limited; more cases are needed for full calibration and external validation. Trans apical TAVI patients were excluded, hence it could be interesting in the future scores performance for this subgroup of patients. In our opinion, observations were limited and data are deriving from single institution.

### Interpretation

EuroSCORE II and ACEF should be avoided for risk stratification in MICS population. STS score could be used, but it has limited application (low to medium risk).

## Conclusions

The STS score is acceptable discrimination power for MICS sub-population. However, it is not calibrated for the the subset. EuroSCORE II is has good discrimination power, but not calibrate for the MICS subset of patients. ACEF score, having only three variables, performs with decent discrimination power but not calibrated for the MICS subset. No algorithm seems suitable for accurate risk estimation in MICS population. Future multicenter analysis is required in order to be able to find most perfomant scoring system. Other scores must be also explored.FiguresPredicted Probability

## Data Availability

Data will be de-identified a will be made available after publication

